# Mapping Power Through Space: The Spatial Intersectionality Health Framework and Intersectional Geographically-Explicit Ecological Momentary Assessment

**DOI:** 10.64898/2026.04.09.26350546

**Authors:** Stephanie H. Cook, Benjamin Pettus

## Abstract

**Background:** Young sexual and gender minorities of color face compound health risks shaped by interlocking systems of racism, cisgenderism, and class inequality. Spatial health research documents that place shapes health, but existing methods cannot specify the mechanisms through which spatial configurations produce different health outcomes for differently positioned people. This gap prevents targeted intervention.

**Objective:** To develop and pilot test the Spatial Intersectionality Health Framework (SIHF), which specifies three mechanisms through which space produces intersectional health inequities: Layered (multiple oppressive systems activating simultaneously), Positional (the same space producing different health pathways by intersectional position), and Conditional (nominally protective spaces carrying hidden costs for specific positions). We also introduce and validate Intersectional Geographically-Explicit Ecological Momentary Assessment (IGEMA) as the methodology operationalizing SIHF across three data levels.

**Methods:** The GeoSense study enrolled 32 young sexual and gender minorities of color (ages 18-29) in New York City. IGEMA was implemented across three integrated levels: (1) GPS mobility tracking via participants’ personal smartphones, linked to census tract structural exposure indices across n=19 participants; (2) ecological momentary assessment of intersectional discrimination with multilevel modeling of mood, stress, and sleep outcomes; and (3) map-guided qualitative interviews with SIHF mechanism coding and intercoder reliability assessment across 92 coded records from 18 participants. This study was conducted as the pilot for NIH R01HL169503.

**Results:** All three SIHF mechanisms were empirically detectable. A compound structural gendered racism index outperformed every single-axis alternative in predicting daily mood (b=-0.048, p=.001) and stress (b=0.121, p<.001). The Positional mechanism accounted for 71% of coded harm experiences. Intercoder reliability for mechanism assignment reached kappa=0.824 at Stage 2 reconciliation. Daily intersectional discrimination predicted greater sleep disturbance (b=1.308, p=.004).

**Conclusions:** SIHF and IGEMA together provide an empirically testable framework for specifying how space produces intersectional health inequities. Mechanism specification, not spatial location alone, is the condition for designing research and intervention that reaches the source of harm for multiply marginalized populations.

## 1. Introduction

A young Black transgender woman named Jasmine moves through her city over the course of a day. In her residential neighborhood, neighbors recognize her and offer familiarity that can feel protective, yet surveillance and gossip circulate through the same social networks. On a crowded subway platform, strangers read her body and respond with curiosity, avoidance, or hostility. In a clinic waiting room, intake forms structured around binary gender categories force her to decide whether to disclose personal information publicly or accept administrative erasure. At work, professional norms regulate how she presents her gender and how colleagues respond to it. Jasmine’s intersectional position does not change as she moves between these spaces, yet the forms of risk, visibility, safety, and belonging she encounters shift dramatically across locations. The question is not only who Jasmine is, but how the systems of power that organize space act on her as she moves through it.

Health inequities research has long established that interlocking systems of oppression shape population health. Intersectionality theory provides the conceptual foundation for understanding why. Emerging from Black feminist thought and activism, intersectionality emphasizes that systems such as racism, sexism, cisgenderism, and class inequality operate simultaneously rather than independently. The Combahee River Collective (1977) articulated this insight in identifying the interlocking nature of major systems of oppression. Crenshaw’s foundational legal scholarship demonstrated how discrimination frameworks designed around single axes of identity fail to capture the experiences of individuals positioned at multiple intersections of power (Crenshaw, 1989, 1991). Sociological scholarship further extended these insights by analyzing how intersecting systems structure social life across institutional and interpersonal domains (Collins, 2000). Over the past several decades, scholars have applied intersectionality across fields including sociology, public health, psychology, and political science, demonstrating that intersecting systems of power shape exposure to discrimination, access to resources, and vulnerability to health risks (Cole, 2009; Bowleg, 2012; Bauer, 2014; Homan et al., 2021).

Despite these advances, intersectionality research has primarily focused on identifying who experiences interlocking oppressions rather than explaining how those experiences unfold across space. Spatial and neighborhood health research demonstrates that where people live and spend time shapes health through environmental conditions, institutional resources, and patterns of social organization (Diez Roux et al., 2001; Krieger, 2001; Massey & Denton, 1993; Sallis et al., 2016; Williams & Collins, 2001). These spatial processes contribute to persistent inequalities in chronic disease, mental health, and life expectancy (Bambra, 2016; Chetty et al., 2016; Macintyre et al., 2002). These approaches have produced important insights into the spatial distribution of health inequities, yet they typically conceptualize space as a container of exposure rather than as a structural axis through which systems of power operate (Diez Roux & Mair, 2010; Arcaya et al., 2012; Kim et al., 2022). Spatial health research can identify where health outcomes cluster, but it often cannot explain why the same space produces different experiences for differently positioned people. A park, clinic, transit station, or workplace may represent safety for some individuals and vulnerability for others. These differences do not arise from space alone or from identity alone. Instead, they emerge from the interaction between spatial configurations and intersectional social positions.

Foundational structural determinants frameworks have made essential contributions to understanding how place shapes health. Krieger’s (2001) ecosocial theory establishes that individuals incorporate the material and social conditions they inhabit, including the physiological consequences of navigating unequal power relations. Link and Phelan’s (1995) fundamental cause theory explains why social inequalities persist across changing disease landscapes by controlling access to protective resources. Gee and Ford’s (2011) structural racism framework identifies residential segregation and neighborhood conditions as downstream pathways through which racism produces health inequities. These frameworks treat place as a level of exposure or a pathway through which upstream social conditions operate. None specifies the mechanisms through which spatial configurations activate interlocking systems differently depending on intersectional position, a problem the spatial intersectionality literature has begun to address.

Building on that scholarship, researchers have established that intersecting systems of power operate through space and that spatial context shapes who experiences what forms of oppression. Forde (2024) documents how Cape Town’s townships concentrate risk on young people facing intersecting oppressions. Morrell and Blackwell (2022) trace how Milwaukee’s segregation patterns concentrated Black women in lead-contaminated neighborhoods. Rodó-de-Zárate (2023) visualizes how the same plaza produces comfort for some and fear for others, depending on intersectional identity position. Bambra (2022) calls for placing intersectional inequalities in health, arguing that place should be considered alongside race, gender, and class as a fundamental dimension in health equity analysis. A broader body of scholarship in feminist geography and Black geography has established that spatial environments are organized through intersecting systems of power and that bodies navigate geographies structured by their intersectional social positions (Gkiouleka et al., 2018; Lipsitz, 2007; Massey, 1994; McKittrick, 2006; Mollett & Faria, 2018; Rodó-de-Zárate and Baylina, 2018; Valentine, 2007).

What the existing literature leaves unresolved is not whether space matters for intersectional health but how it operates to produce health inequities. Prior work identifies the phenomenon without specifying the mechanism. Knowing that the same place produces different health experiences for differently positioned people does not explain why, and knowing that space and interlocking systems of oppression are co-constitutive does not tell us what that relationship does at a specific moment for a specific person. Describing the phenomenon is not the same as specifying the mechanism. The most direct methodological predecessor to Intersectional Geographically-Explicit Ecological Momentary Assessment (IGEMA) is Rodó-de-Zárate’s Relief Maps methodology (2014, 2023). Relief Maps is a three-dimensional approach that captures social positions, places, and emotional experience simultaneously. It visualizes how the same location, such as a plaza, a clinic, or a transit stop, produces ease for some people and oppression for others, depending on their intersectional identity position. It establishes identity-space interaction as the unit of analysis and uses map-guided interviews as its primary data collection tool, asking participants to narrate their experiences at specific locations they have moved through. The methodology builds directly on this design. The GPS trace functions as Rodó-de-Zárate’s elicitation device, grounding participant accounts in the specific spatial encounters of their monitored days rather than general recollection.

Relief Maps documents that variation exists, but variation alone does not describe a mechanism. Documenting that the same clinic produces differential experiences for people at different intersectional positions is the observation. Documenting that a wheelchair-accessible entrance routed through a service corridor simultaneously exposes a Black wheelchair user to racialized surveillance in that low-visibility route, and ableist exclusion through the segregated entry is the observation. Without knowing which mechanism is producing those differences, researchers cannot identify what to change, clinicians cannot know where to intervene, and policymakers cannot design reforms that reach the source of harm rather than its most visible surface. The framework specifies what is producing the disparity by assigning one or more of three distinct mechanisms. Whether a space produces differential health effects because it is 1) layered, meaning multiple systems are activated simultaneously; 2) positional, meaning it was organized around one intersectional position’s needs and not others; or 3) conditional, meaning a protective design carried hidden costs for specific positions. These are three different explanations with three different implications for health intervention. Without naming those modes, researchers cannot empirically distinguish among them or design interventions that target the right pathway.

We build on this previous work in spatial intersectionality by introducing SIHF, a framework that conceptualizes space as a constitutive structural axis through which interlocking systems of oppression produce health inequities. Spaces do not simply trigger systems of power when a person enters them. They are produced through those systems, often organized around an assumed occupant whose intersectional position differs from those of multiply marginalized people. Layered operates when multiple systems of oppression activate simultaneously through spatial configurations; Positional, in which the same spatial environment produces qualitatively different experiences depending on intersectional social position; and Conditional, in which spaces designed to provide protection or inclusion produce unintended harms for particular intersectional groups. To operationalize this framework empirically, the paper introduces IGEMA, which integrates GPS-linked structural exposure measures, ecological momentary assessments of discrimination and health outcomes, and map-guided qualitative interviews to detect and distinguish among mechanisms in real time (McQuoid et al., 2018). Together, SIHF and IGEMA provide the theoretical and methodological architecture for explaining how spatial environments structure the activation of interlocking systems of oppression and for detecting those mechanisms empirically in health research. Using pilot data from the GeoSense study of young sexual and gender minorities of color in New York City, we demonstrate how this integrated design enables researchers to detect spatial intersectional mechanisms that would remain invisible within single-method approaches.

## 2. The Spatial Intersectionality Health Framework

The Spatial Intersectionality Health Framework, or SIHF, conceptualizes space as a structural axis through which interlocking systems of oppression shape health inequities. Rather than treating spatial context as a background condition, SIHF focuses on how spatial environments organize the interaction of intersecting systems of power during everyday life. The framework shifts analytic attention from where inequality occurs to how specific spatial configurations activate intersecting forms of marginalization.

The framework identifies three mechanisms through which spatial dynamics operate. The Layered mechanism describes situations in which multiple systems of oppression become active within the same spatial configuration. The Positional mechanism captures how the same space produces different experiences depending on an individual’s intersectional social position. The Conditional mechanism identifies cases in which spaces designed to address inequality generate unintended harms when they assume a simplified or normative user. Together, these mechanisms specify how spatial environments translate structural inequality into lived experience and ultimately into health consequences (Figure 1). The framework also generates the concept of spatial concordance, the condition in which the structural composition of a space aligns with a person’s intersectional position, reducing identity-mismatch activation and generating relief rather than harm.

**Figure 1.**
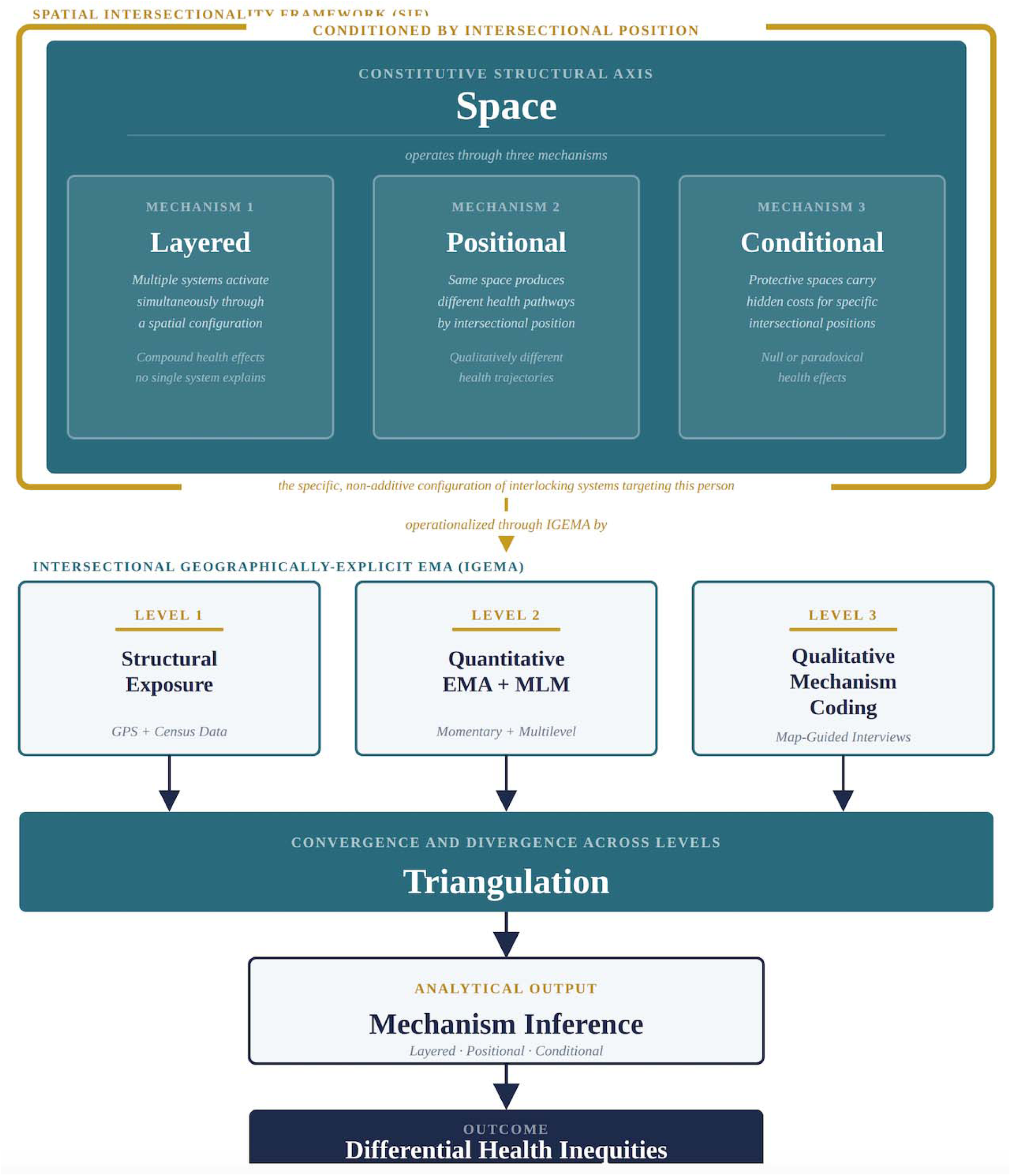
The Spatial Intersectionality Health Framework and Its Operationalization Through IGEMA. The Spatial Intersectionality Health Framework and Its Operationalization Through IGEMA. The upper section presents the Spatial Intersectionality Health Framework (SIHF), in which space operates as a constitutive structural axis producing differential health inequities through three mechanisms: Layered (simultaneous activation of multiple systems through a spatial configuration), Positional (the same space producing qualitatively different health pathways depending on intersectional position), and Conditional (nominally protective spaces carrying structurally predictable hidden costs at specific intersectional positions). The gold frame represents intersectional position, the specific non-additive configuration of interlocking systems targeting a given person, which conditions all three mechanisms. A bridge arrow connects SIHF to the lower section, which presents the IGEMA architecture. IGEMA operationalizes all three mechanisms through three integrated measurement levels: Level 1 (GPS-linked structural exposure data) establishes where each mechanism is structurally predicted to be active; Level 2 (ecological momentary assessment and multilevel modeling) tests whether the health effects each mechanism predicts are detectable in the data; and Level 3 (map-guided qualitative interviews and mechanism coding) identifies which systems were operating at specific locations for specific intersectional positions. Triangulation across all three levels produces mechanism inference, specifying which mechanism organized differential health inequities at each location.

### 2.1 Layered Mechanism

The Layered mechanism describes situations in which multiple systems of oppression activate simultaneously through the same spatial configuration. Harm does not result from one axis of inequality acting alone. Instead, intersecting structures such as racism, sexism, cisgenderism, and class inequality converge within a specific environment to produce interlocking forms of marginalization. The mechanism is intersectional because health consequences emerge from the interaction among systems of power rather than from the additive presence of separate disadvantages.

Healthcare settings illustrate this dynamic clearly. Many clinical spaces are organized around assumptions about normative patients who are implicitly White, cisgender, able-bodied, and economically stable. Administrative routines such as binary intake forms, open waiting rooms, and public name-calling practices can force disclosure, misrecognize gendered identities, and expose patients to scrutiny in highly visible settings. For transgender patients of color, these institutional arrangements may also activate racialized assumptions about credibility, danger, or economic status. Racism and cisgenderism, therefore, become jointly operational through the spatial organization of the clinic itself (Sevelius, 2013; Poteat et al., 2013). A clinic organized around assumptions of a normative White cisgender patient activates racism and cisgenderism simultaneously through the same spatial encounter. In an open waiting room, a Black transgender patient navigates racialized surveillance from staff and other patients at the same moment their legal name is called aloud, forcing gender disclosure in a public setting. In the exam room, racialized patterns of pain dismissal and cisgenderist assumptions about anatomy compound through the same provider interaction. Neither system requires the other to produce harm, but both activate through the same spatial configuration (Institute of Medicine, 2011; Balsam et al., 2011).

Layered mechanisms also operate at broader structural scales. Residential segregation concentrates environmental hazards, economic deprivation, and institutional disinvestment within the same neighborhoods while simultaneously shaping access to healthcare, transportation, healthy food environments, and political power (Bullard, 1990; Massey & Denton, 1993; Morello-Frosch et al., 2011; Williams & Collins, 2001). These exposures reinforce one another through spatial arrangements that reproduce racialized and class inequality over time, producing health outcomes such as chronic stress exposure, cardiometabolic risk, and reduced life expectancy through their intersectional concentration (Diez Roux & Mair, 2010; White & Borrell, 2011; German & Latkin, 2012).

### 2.2 Positional Mechanism

The Positional mechanism describes situations in which the same spatial environment produces different experiences depending on an individual’s intersectional social position. The physical setting remains constant, yet its social meaning shifts because systems of power structure how bodies are interpreted within it. Spaces carry histories of racialized and gendered exclusion that shape who belongs and who is subject to surveillance regardless of formal changes to access (Lipsitz, 2007; Massey, 1994; McKittrick, 2006). This mechanism highlights that spatial environments do not have uniform effects independent of the people who occupy them.

Public parks provide a useful illustration. Research on gendered racism and racialized surveillance demonstrates that public environments are rarely experienced as neutral (Essed, 1991; Wingfield, 2007). Black men in particular are frequently subject to heightened scrutiny because cultural stereotypes associate Black masculinity with danger or criminality, and spatial features such as open sightlines and police patrol routes intensify these dynamics (Fagan & Geller, 2015; Goff et al., 2014; Welch, 2007). A Black man sitting on a bench in a predominantly White neighborhood may attract attention from residents or police officers who interpret the same behavior as suspicious. A White woman in the same setting may be read as belonging even as she navigates gendered concerns about safety (Valentine, 1989; Pain, 1997). A Black woman may encounter a different set of dynamics shaped by the intersection of racism and sexism (Collins, 2004; Crenshaw, 1991). The spatial environment remains constant, yet the interaction between place and intersectional position produces fundamentally different experiences. Repeated exposure to vigilance, scrutiny, or exclusion produces chronic physiological stress and hypervigilance that accumulates into health consequences over time (Carter, 2007; Pieterse et al., 2012).

### 2.3 Conditional Mechanism

The Conditional mechanism describes situations in which spaces designed to protect or benefit marginalized populations generate unintended harms for individuals at particular intersections of identity. These harms arise when spatial interventions address one dimension of inequality while leaving other systems of power unexamined. Inclusion defined along a single axis may therefore reproduce exclusion for people whose experiences are shaped by multiple intersecting positions.

Urban greening initiatives illustrate this dynamic. Investments in parks, tree canopy, and recreational infrastructure are often implemented in historically disinvested neighborhoods of color to address environmental justice and improve public health (Bullard, 1990; Wolch et al., 2014; Jennings et al., 2016). However, new parks and environmental amenities can also increase neighborhood desirability and property values. Rising housing costs may displace long-term residents who already face racial and economic marginalization, disrupting housing stability, social networks, and access to community institutions that support health (Anguelovski et al., 2019; Fullilove, 2004). The intervention was intended to improve environmental conditions, but its benefits remain conditional on residents’ ability to remain in place.

### 2.4 Mechanisms in Concert

The Layered, Positional, and Conditional mechanisms are analytically distinct, but they often operate together within the same spatial encounter. Consider a wheelchair-accessible entrance located along a building service corridor. The design may activate the Conditional mechanism because accessibility is treated as an accommodation rather than a fully integrated design principle (Imrie, 1996; Boys, 2014). At the same time, the Positional mechanism may operate if racialized bodies encounter heightened surveillance within that low-visibility route (Goff et al., 2014). If ableism and racism are activated simultaneously through the same setting, the encounter may also reflect a Layered mechanism. One spatial configuration can therefore organize multiple interlocking systems of power at once.

Mechanisms also shift in response to intervention. An LGBTQ-affirming clinic designed to address cisgenderism may substantially reduce harm along the gender identity axis while leaving race unexamined. For a queer patient of color, the space has moved from Positional to Conditional: it is now nominally protective, but that protection is partial, and the unprotected racial axis becomes the site of harm. Designing for one axis while leaving others structurally unaddressed does not eliminate the mechanism. It changes which one is operating.

Recognizing how these mechanisms interact is essential for understanding how spatial environments generate intersectional health inequities. People moving through cities encounter overlapping spatial processes that shape everyday experiences of safety, stress, belonging, exclusion, and recovery. Detecting these mechanisms requires methods capable of linking spatial context, lived experience, and health outcomes as they unfold during daily life. The following section introduces IGEMA as the methodological architecture used to capture these dynamics empirically.

## 3. Intersectional Geographically Explicit Ecological Momentary Assessment

### 3.1 From Spatial Behavior to Spatial Power: Why Existing Methods Are Insufficient

The theoretical architecture of SIHF creates a specific methodological demand. If space operates as a structural axis through which interlocking systems of oppression produce health inequities, then methods designed to study space as context, background, or behavioral setting cannot test that claim. Existing residential neighborhood approaches assign individuals to fixed administrative units and treat the neighborhood as a container for exposures, measuring where people live rather than the specific places they occupy across a day (Arcaya et al., 2012; Cummins et al., 2007). Standard ecological momentary assessment captures health experiences as they occur but without spatial specificity (Bolger & Laurenceau, 2013), providing no way to test whether the same spatial configuration activated a different mechanism for someone at a different intersectional position.

Geographically Explicit Ecological Momentary Assessment, established in health research by McQuoid, Thrul, and Ling (2018), addressed part of this problem by linking momentary assessments to GPS-recorded locations. The methodology is extended here by reorienting the theoretical target. GEMA locates behavior in space. The present framework locates the operation of power through space. The distinction is not methodological refinement. It is a different analytical object. GEMA asks what situational factors at a location predict a behavior. The present framework asks which interlocking systems of oppression are structurally active at a location, whether power acted through those systems on a person whose intersectional position made them a target, and what health effect resulted. The methodology also builds directly on Rodó-de-Zárate’s Relief Maps methodology, which established identity-space interaction as the unit of analysis and used map-guided interviews to document how the same location produces different experiences depending on intersectional position. Three extensions distinguish IGEMA from Relief Maps: GPS-recorded traces replace self-selected maps, grounding accounts in the specific spatial encounters of participants’ monitored days; each location is linked to census-derived structural indicators so participant accounts can be triangulated against structural predictions; and the qualitative interview is embedded within a three-level architecture designed to produce mechanism inference rather than experiential documentation alone. Relief Maps is the methodological origin of the identity-space interview. The present methodology is its extension into mechanism detection. Full specification of the IGEMA design and codebook protocol is provided in Supplement Notes S1 and S2. (Figure 1)

### 3.2 The IGEMA architecture

Mechanism inference is the core analytic output of IGEMA, a determination of which interlocking systems of oppression were active at a specific location, through which of the three SIHF mechanisms, producing which health effect, for a person whose intersectional position made them a target. No single data source produces that inference. Structural data alone establishes spatial conditions but cannot confirm that power acted on a particular person at a particular moment. Quantitative models alone can detect health effects, but cannot identify the mechanism or the intersecting systems responsible. Qualitative accounts alone can name what was operating, but cannot confirm the structural conditions that organized that operation or test whether the health effect was detectable across the sample. Mechanism inference requires all three, triangulated.

The three evidentiary layers each answer a different question. The first asks where each SIHF mechanism is structurally predicted to be active. GPS monitoring captures movement through activity spaces (Chaix et al., 2012), and each visited location is linked to census-derived structural indicators indexing the spatial concentration of racial, economic, and sexual minority conditions at that place (Krieger et al., 2016; Homan et al., 2021). The second asks whether the health effect each mechanism predicts is detectable in the data? Multilevel models separate within-person from between-person structural effects, distinguishing acute activation at specific locations from chronic structural burden accumulated across repeated spatial navigation (Bolger & Laurenceau, 2013). The third asks, which systems were actually operating at this location for this person? Map-guided qualitative interviews, conducted after the monitoring period with the GPS record as an elicitation device, ground participant accounts in the specific spatial encounters of the monitored days rather than general experience (Rodó-de-Zárate, 2014; McQuoid et al., 2018). Coders assign each GPS-linked location a space type, an active intersection code, and a spatial mechanism code. The mechanism code is the finding; space type and intersection are the evidentiary inputs that make mechanism assignment possible.

Triangulation across the three layers proceeds through convergence and divergence. Convergence, where a location is characterized as a site of harm at high structural exposure, and the model detects the compound health effect that the Layered mechanism predicts, provides strong evidence of Layered activation. Divergence, where a site is characterized as a site of harm at a location where structural indicators predicted safety, identifies Positional dynamics that aggregate structural measures cannot detect. In IGEMA, the integration is the method. The inferential target, mechanism activation, is constructed across all three layers simultaneously, and the meaning of any single layer’s evidence depends on what the others show.

### 3.3 Pilot application: the GeoSense study

The GeoSense pilot study was designed to test whether SIHF’s three mechanisms are detectable in real-world data, whether IGEMA’s three evidentiary layers converge and diverge in theoretically predicted patterns, and whether integrated mechanism verdicts emerge that no single layer would produce independently. The pilot is a proof-of-concept for the inferential architecture, not a study designed to estimate the prevalence or magnitude of any mechanism’s effects.

The GeoSense pilot study recruited 32 young sexual and gender minorities of color (ages 18-29) in New York City. The GPS subsample (n = 19) was broadly representative of the full analytic sample demographically. Participants completed a 7-day protocol combining continuous GPS monitoring via Google Maps location history, actigraphy (ActiGraph GT9X), eight daily EMA assessments, a nightly diary, and a map-guided qualitative interview conducted within one week of the monitoring period. Participants enabled location tracking on their personal smartphones throughout the monitoring period and shared their Google Maps Timeline data file with the study team at completion (Fowler, 2023). Individualized maps displaying GPS traces and Level 1 structural exposure data were generated before each qualitative interview, grounding accounts in specific spatial encounters across the monitoring period. Sample characteristics are reported in Table 1. Level 1 structural exposure summary statistics are reported in Table 2. Full protocol detail is in Supplement Note S2.

**Table 1.** GeoSense Pilot Study Sample Characteristics.

| Variable | Full sample N = 32 | GPS subsample n = 19 | Qualitative subsample n = 18 |
| --- | --- | --- | --- |
| **Age** |  |  |  |
| 18–24 | 22 (68.8) | 15 (78.9) | 15 (83.3) |
| 25–29 | 10 (31.3) | 4 (21.1) | 3 (16.7) |
| **Race/ethnicity** |  |  |  |
| Hispanic/Latino | 11 (34.4) | 5 (26.3) | 5 (27.8) |
| Asian/Pacific Islander | 8 (25.0) | 3 (15.8) | 3 (16.7) |
| White | 6 (18.8) | 5 (26.3) | 4 (22.2) |
| Black/African American | 4 (12.5) | 3 (15.8) | 3 (16.7) |
| Multiracial | 2 (6.3) | 2 (10.5) | 2 (11.1) |
| Other | 1 (3.1) | 1 (5.3) | 1 (5.6) |
| **Gender identity** |  |  |  |
| Cisgender Woman | 17 (53.1) | 11 (57.9) | 11 (61.1) |
| Non-binary/GQ/GNC | 5 (15.6) | 3 (15.8) | 2 (11.1) |
| Cisgender Man | 4 (12.5) | 3 (15.8) | 3 (16.7) |
| Transgender Woman | 2 (6.3) | 2 (10.5) | 2 (11.1) |
| Other/Agender | 3 (9.4) | 0 (0.0) | 0 (0.0) |
| Transgender Man | 1 (3.1) | 0 (0.0) | 0 (0.0) |
| **Sexual orientation** |  |  |  |
| Bisexual | 16 (50.0) | 10 (52.6) | 10 (55.6) |
| Other | 7 (21.9) | 5 (26.3) | 5 (27.8) |
| Lesbian | 5 (15.6) | 2 (10.5) | 2 (11.1) |
| Gay | 4 (12.5) | 2 (10.5) | 1 (5.6) |
| **Education (n = 31, 18, 17)<sup>a</sup>** |  |  |  |
| College degree | 18 (58.1) | 10 (55.6) | 9 (52.9) |
| Graduate degree | 7 (22.6) | 4 (22.2) | 4 (23.5) |

| Variable | Full sampleN = 32 | GPS subsamplen = 19 | Qualitative subsamplen = 18 |
| --- | --- | --- | --- |
| Some college | 5 (16.1) | 4 (22.2) | 4 (23.5) |
| Grade school | 1 (3.2) | 0 (0.0) | 0 (0.0) |
| **Annual income (n = 28, 15, 14)a** |  |  |  |
| \$0–\$10,999 | 16 (57.1) | 9 (60.0) | 9 (64.3) |
| \$11,000–\$20,999 | 4 (14.3) | 2 (13.3) | 1 (7.1) |
| \$31,000–\$40,999 | 2 (7.1) | 1 (6.7) | 1 (7.1) |
| \$51,000–\$60,999 | 4 (14.3) | 2 (13.3) | 2 (14.3) |
| Prefer not to answer | 2 (7.1) | 1 (6.7) | 1 (7.1) |
| **Employment status (n = 31, 18, 17)a** |  |  |  |
| Working | 16 (51.6) | 10 (55.6) | 9 (52.9) |
| Unemployed | 12 (38.7) | 7 (38.9) | 7 (41.2) |
| Other/Student | 3 (9.7) | 1 (5.6) | 1 (5.9) |

*Note.* Values are n (%). GQ = Genderqueer; GNC = Gender non-conforming.
*a* Missing responses excluded from percentage calculations.
*GPS subsample* = participants who completed continuous GPS monitoring via Google Maps location history. *Qualitative subsample* = GPS participants with completed map-guided qualitative interview and Level 3 coding (excludes one participant who completed GPS monitoring but not the qualitative interview).

**Table 2.** Level 1 Structural Exposure: Sample-Level Summary (GPS Subsample, n = 19)

| Indicator | Sample mean (SD) | Range |
| --- | --- | --- |
| **Racial spatial polarization** |  |  |
| Racial structural exposure (%) | 36.02 (27.14) | 3.80 to 93.20 |
| ICE_race (Index of Concentration at Extremes) | -0.03 (0.43) | -0.64 to 0.74 |
| **Economic deprivation** |  |  |
| Economic structural exposure (%) | 12.69 (6.17) | 0.70 to 25.50 |
| ICE_income | -0.07 (0.16) | -0.35 to 0.31 |
| **Compound structural gendered racism** |  |  |
| ICE_gr (compound index) | -0.05 (0.28) | -0.49 to 0.52 |
| **Structural power quadrant distribution (home tract)** |  |  |
| Concentrated Structural Disadvantage (CSD) | n = 3 | 15.8% |
| Racialized Polarization (RP) | n = 2 | 10.5% |
| Economic Deprivation (ED) | n = 3 | 15.8% |
| Structural Advantage (SA) | n = 11 | 57.9% |

*Note. Values are person-level means (one observation per GPS participant), averaged across all monitored days. ICE = Index of Concentration at Extremes (Krieger et al., 2016). ICE\_race: proportion White non-Hispanic minus proportion Black non-Hispanic at census tract level; ICE\_income: proportion high-income minus proportion low-income households. ICE\_gr = compound structural gendered racism index (mean of ICE\_race and ICE\_income). Racial structural exposure: % Black non-Hispanic + Hispanic/Latino residents in home census tract. Economic structural exposure: mean of tract poverty rate and unemployment rate. Structural power quadrant thresholds: racialized polarization $\geq 50\%$ ; concentrated economic deprivation $\geq 15\%$ (following Krieger et al., 2016).*

GPS traces were linked to census tract-level structural indicators from the American Community Survey (ACS 2017-2021 5-year estimates), capturing racialized spatial polarization, concentrated economic deprivation, and structural exposure to sexuality. A compound index of structural gendered racism (SGR; operationalized as ICE_gr), used as a proxy for the simultaneous concentration of racial and economic spatial disadvantage (Krieger et al., 2016), encoded this compound formation. Within-person and between-person decomposition followed multilevel centering procedures (Bolger & Laurenceau, 2013). Intersectional discrimination was assessed using the Intersectional Discrimination Index (InDI; Scheim & Bauer, 2019). Three-level multilevel models (observations within days within persons) estimated within-person and between-person associations between discrimination, structural exposure, and health outcomes across three outcomes, mood, stress, and sleep disturbance. Before running intersectional specifications, within-person correlations among structural predictors were examined; within-person economic structural exposure and SGR were correlated at *r* = 0.848, and accordingly, within-person economic exposure was excluded from all specifications containing SGR, while between-person economic exposure was retained as a contextual control. Full model output is in Supplement Table S2. Key model comparison results are reported in Table 3.

**Table 3.** Level 2 Key Results: ICE_gr Model Comparisons (GPS Subsample, n = 15)

| Outcome | Within-person b (p) | Between-person b (p) | R<sup>2</sup> discrim.-only | R<sup>2</sup> ICE_gr model | LRT $\chi^2$ (p) |
| --- | --- | --- | --- | --- | --- |
| Mood | 26.75 (.008) | -27.62 (.003) | 0.033 | 0.472 | $\chi^2(6)=17.5$ (.007) |
| Stress | -33.33 (.002) | 29.34 (.003) | 0.032 | 0.318 | $\chi^2(6)=18.9$ (.004) |
| Sleep disturbance | n.s. | n.s. | 0.034 | 0.211a | $\chi^2(4)=14.39$ (.006) |

*Note.* *b* = unstandardized coefficient from three-level multilevel model. *R*<sup>2</sup> = marginal *R*<sup>2</sup> (Nakagawa & Schielzeth, 2013). *LRT* = likelihood ratio test vs. discrimination-only baseline. *ICE\_gr* = compound structural gendered racism index. *a* Race-by-economic specification. *n.s.* = not significant in *ICE\_gr* specification.

Map-guided qualitative interviews were audio-recorded, professionally transcribed, and linked to GPS records by participant identifier and monitoring day (Rodó-de-Zárate, 2014; McQuoid et al., 2018). Two independent coders evaluated each GPS-linked excerpt to assign space type, active intersections, and spatial mechanism. Classification rules, decision procedures, and codebook development are detailed in Supplement Note S1. Pre-calibration intercoder agreement met the pre-specified κ ≥ 0.70 threshold for space_type (κ = 0.68–0.70) but fell below the threshold for spatial mechanism (κ = 0.54; Krippendorff, 2004), with disagreements concentrated on Layered versus Conditional and Layered versus Positional boundaries. A structured calibration session resolved these boundary conditions through decision rule clarification and anchor case review. Post-calibration agreement exceeded thresholds for both codes (space_type κ = 1.00; mechanism κ = 0.80; Cohen, 1960; Landis & Koch, 1977) across 91 coded records. Full reliability statistics are in Table S4.

#### Triangulation: What the Three Levels Show Together

The three levels converge on a finding that none produce independently. For young SGM of color in New York City, spatial inequities are organized through intersectional position, not primarily through structural disadvantage at the location.

The Level 2 results present two interpretive puzzles that no single measurement layer can resolve on its own. The first is that SGR predicted opposite effects depending on whether it was capturing a person’s daily movements or their typical home environment. On days when participants moved through tracts with higher SGR concentrations than their usual pattern, they reported better mood (*b* = 26.75, *p* = .008) and lower acute stress (*b* = −33.33, *p* = .002). Yet participants whose typical neighborhoods carried higher average SGR concentrations reported chronically lower mood (*b* = −27.62, *p* = .003) and higher baseline stress (*b* = 29.34, *p* = .003) across all monitoring days. Being near structural gendered racism on a given day was protective for most participants. Living inside it over time was not. Level 3 explains why. The second puzzle is outcome specificity. The structural gendered racism index that strongly predicted mood and stress did not predict sleep disturbance, while the race-by-economic structural specification did (*χ²*(4) = 14.39, *p* = .006, R²m 0.034 → 0.211 (Nakagawa & Schielzeth, 2013). Level 3 qualitative data and the spatial logic of Level 1 together explain both patterns.

The within-person divergence is a home story. The tracts with the highest SGR concentrations in this sample are predominantly home neighborhoods. Days when participants occupied those tracts were days closer to home, with less cross-boundary movement into the institutional settings (i.e., workplaces, campuses, healthcare facilities, commercial environments, transit) where the Level 3 data shows inequities concentrating. The within-person protective pattern does not reflect a direct health benefit of structural disadvantage. It reflects the spatial rhythm of home proximity, which for most participants means proximity to racial and ethnic concordance, protection from identity-mismatch activation, and reduced daily exposure to the positional encounters that produce acute stress and affective disruption. The between-person pattern runs in the opposite direction for the same structural reason. Chronic residence in a neighborhood of concentrated gendered racial disadvantage is an accumulating burden that shapes the baseline physiological state a participant brings to every monitoring day, regardless of daily spatial movements.

The Level 3 data shows this logic in individual accounts. Sofia, a Latina cisgender bisexual woman living in Spanish Harlem, described her home neighborhood as the place she fit in better than anywhere else, which is a source of racial and ethnic belonging that produced daily ease. For Sofia, the neighborhood’s racial and ethnic composition aligned with her identity in a way that buffered rather than threatened. Her workplace produced a different pattern entirely. Working remotely for a predominantly White organization, she described having to work harder to prove she deserved to be there, naming both her identity and her qualifications as simultaneous dimensions of competition. “I feel like I have to work a little bit harder to prove that I deserve to be here… not only competing because of my identity but also qualifications. I don’t usually say something because I know they mean well.” According to SIHF, this is the signature of the layered mechanism, race and class locking together in a single institutional setting such that neither axis alone accounts for the intersectional burden.

The within-person buffering that most participants accessed through home proximity was not uniformly available. Jordan, a Hispanic and Latino non-binary person in the South Bronx, whose home neighborhood carries the highest SGR concentration in the sample, described their home area as a source of harm rather than relief. They described feeling like they were sticking out in their own neighborhood, with people following closely behind them, and having to change where they walked to manage the exposure. They had cut their hair and dressed differently and observed that changes in their gender presentation altered how they were treated in the immediate neighborhood. To find ease, they had to travel. “I will tend to venture out into the city to be around people who are my age, my demographic, to feel a bit more at ease with myself.” When a person’s intersectional identity formation does not match the prevailing norms of their neighborhood, high SGR proximity produces harm rather than relief. This is the positional mechanism at work and exemplifies the condition under which the aggregate within-person protective pattern reverses.

Level 3 also shows that structural disadvantage is not the only condition under which harm operates. Across the sample, oppression was coded at commercial retail environments, institutional settings with binary gender organization, and street-level encounters, regardless of the structural profile of the surrounding tract. Mei, an Asian Pacific Islander transgender woman, described a retail visit in which she was misgendered by a store employee as “a huge” disruption, and described a Sephora visit as “really stress-inducing because there’s like a lot of cis women in there,” noting that even compliments felt marked, as if people “say that because they feel the need to because I’m trans.” Both locations were in structurally mixed or advantaged tracts. The positional mechanism, which accounted for the majority of coded harm experiences across the sample, activates through the relationship between the participant’s intersectional position and the space’s assumed occupant, not through the neighborhood’s structural disadvantage level. That is why single-axis structural specifications failed to improve model fit in the Level 2 analysis while the SGR index succeeded: the compound measure encodes the gendered racial power differential that organizes institutional spaces, not only the neighborhood poverty rate.

Level 3 further shows that protection along one identity axis does not extend to others. Simone, a Black/African American cisgender woman with Other orientation, described a community context in which racial identity was protective, but gender and body were not. “I’m more of an object in my community. I feel more discriminated against based on my body type and my gender, but not so much my race or sexuality.” The conditional mechanism is visible here. The space was organized around a shared racial and ethnic identity that functioned as genuinely protective, while the prevailing norms left gender and body entirely unaddressed, exposing those dimensions of her intersectional formation to harm. The SGR index does not capture this. It measures the tract-level concentration of gendered racial disadvantage relative to the dominant group; it does not measure whether community spaces within that tract enforce gender or body norms that harm specific participants. Her spatial experience of harm is not predicted by the structural index because the harm does not arise from her general intersectional mismatch with the space, which would be the basis for Positional coding, but from the prevailing norms that racial concordance is sufficient protection, leaving other dimensions of her intersectional formation structurally unaddressed. That is the Conditional signature.

The sleep divergence from the mood and stress pattern follows a different logic. The structural gendered racism index captures daily variation in identity-mismatch exposure. This produces an acute, event-proximate pathway to affective disruption. Economic structural exposure indexes something more chronic and materially immediate, measuring the concentration of poverty, resource scarcity, and neighborhood deprivation that participants live inside rather than move through. The race-by-economic specification predicted sleep disturbance because sleep disruption accumulates from sustained physiological arousal in environments where material deprivation requires ongoing vigilance, and where the body does not return to baseline between exposures. The SGR index measures daily identity-mismatch activation, not residential material conditions. The two capture distinct spatial processes with distinct health consequences, and the outcome-specific pattern in the Level 2 data reflects that distinction.

The three levels together produce an account that no single measurement approach could generate alone. The divergent within-person and between-person effects of SGR on affect are not statistical artifacts. They reflect a spatial system in which home proximity buffers acute identity-mismatch activation for most participants while chronic residence under structural gendered disadvantage accumulates as baseline burden, and in which that same home can be a site of harm for participants whose intersectional identity formation does not match its prevailing norms. Taken together, the three levels make this visible. Any one of them alone does not.

The pilot was designed to demonstrate that all three SIHF mechanisms are detectable in real-world data, that IGEMA’s three layers converge and diverge in the theoretically predicted patterns, and that integrated mechanism verdicts emerge that no single layer would produce independently. The 19-participant GPS sample accomplishes these proof-of-concept goals. It is not designed to produce stable estimates of mechanism prevalence or effect size magnitude. The sexuality structural exposure index was assigned at the county level due to the ACS suppression of tract-level same-sex household data, limiting its contribution to within-itinerary mechanism detection. Map-guided interviews use GPS traces as elicitation prompts, which may direct participant attention toward researcher-highlighted locations; the protocol addresses this through open-ended prompting. All three considerations specify the interpretive frame within which Level 3 accounts should be read and do not undermine mechanism inference, as Level 3 is one of three independent bodies of evidence and is never the sole basis for a mechanism verdict. Together, the three levels demonstrate what the pilot was designed to show. Mechanism inference is possible, the evidentiary layers converge and diverge in theoretically predicted patterns, and the integrated finding is not recoverable from any single method.

## 4. Discussion

Introduced at the outset of this paper, Jasmine’s experience navigating through a day in her life in New York City raises an important question. How are the spaces she inhabits organized by intersecting systems of power, and what do those configurations do to her health at each location? While observing the same person throughout the same day, the mechanisms producing harm and relief shifted at every location she entered. Intersectionality theory established that people in her position carry health risks that no single-axis framework captures. We also knew that where she lives shapes her health. What we did not have was an architecture for specifying how the systems targeting her activate differently depending on which space she enters, and what that does to her body in real time. That is the gap this paper fills. The Spatial Intersectionality Health Framework names the process. IGEMA provides the method to measure it.

### 4.1 Advancing Intersectionality Through Spatial Mechanisms

The framework contributes to intersectionality scholarship by specifying how spatial environments organize the activation of interlocking systems of racial, gender, sexual, and class oppression for people whose intersectional identity formations make them targets of those systems simultaneously. Intersectionality theory holds that these intersectional formations are qualitatively irreducible to their parts. Gendered racism is not racism plus sexism. Its health effects cannot be recovered by measuring each system separately. The structural intersectionality measurement literature, developed by Homan, Brown, and colleagues (Evans et al., 2018; Homan et al., 2021), confirmed this claim at the population level by showing that compound structural conditions jointly shape health in ways no single-axis measure captures. What SIHF and IGEMA add is the spatial and temporal dimension of that claim. A measure encoding the simultaneous concentration of Black and Latina women in poverty relative to White non-Hispanic men above poverty consistently outperformed every single-axis alternative in predicting the daily mood and stress of young sexual and gender minorities of color, not because race and economic conditions are irrelevant individually but because their intersectional formation captures the specific configuration of power that organizes the spaces these participants moved through each day. Furthermore, within-person and between-person effects of structural gendered racism ran in opposite directions, revealing that the same structural condition acts on the bodies of multiply marginalized people through two distinct temporal mechanisms simultaneously. For most participants, days closer to home were days of reduced activation, home neighborhoods carried racial and ethnic concordance that buffered daily encounters with identity-mismatch. That is not the same as safety from structural burden. Chronic residence under concentrated gendered racial disadvantage shaped baseline stress regardless of where participants traveled on any given day.

Prior scholarship across critical geography, Black feminist geography, and feminist spatial scholarship established that space and intersectional experience are co-constitutive and that the same place produces qualitatively different experiences for people at different intersectional positions (Massey, 1994; McKittrick, 2006; Lipsitz, 2007; Mollett & Faria, 2018). Rodó-de-Zárate’s Relief Maps framework (Rodó-de-Zárate, 2014) made that variation visible and documented for people navigating geographies organized by intersecting systems of power. What remained was the capacity to specify the mechanism driving that variation, to test whether it produces detectable health consequences, and to do both in real time at the level of specific places. The three-level IGEMA design provides that capacity. What prior scholarship described as co-constitutive and irreducible is, it turns out, also measurable.

### 4.2 Methodological Contributions of IGEMA

Two patterns are visible in these findings that existing methods built for other purposes could not detect, precisely because those methods were not designed to observe how spatial power acts on people with intersecting marginalized identities. The first is outcome specificity. The SGR index that strongly predicted daily mood and stress among multiply marginalized young people did not predict sleep disturbance. Sleep was predicted instead by the race-by-economic structural specification. This is not a null result. It reveals two distinct routes through which space acts on the body. The SGR index tracks what happens when a person moves through spaces whose social organization treats their intersectional identity as out of place. That produces an acute stress response on that day, a disruption that people register in their mood and stress levels in real time. Economic structural exposure tracks something different. Living in a neighborhood where material resources are scarce requires a kind of ongoing alertness that does not switch off between days. That sustained physiological state accumulates into disrupted sleep. The SGR index was built to capture daily identity-mismatch activation. The race-by-economic specification captured residential accumulation. Each reached a different health outcome through a different route, and those routes would be invisible without a three-level design.

The second pattern is the dominance of the positional mechanism. Across 91 coded location records, 68% of coded harm experiences (n=32) operated through identity-space mismatch in the institutional settings of daily life, in workplaces, retail environments, transit, and healthcare spaces organized around assumed occupants whose intersectional formations differed from those of the young SGM of color in this sample. Structural concentration of disadvantage at the neighborhood level predicted health outcomes, but it did not account for most of where harm was actually happening to these participants. Approaches that address neighborhood structural conditions alone will not reach the source of most harm in these communities’ daily spatial lives. This matters directly for how interventions are designed. Improving conditions in the neighborhoods where people live addresses a real and cumulative burden. But for the participants in this study, most spatial harm was happening in workplaces, transit systems, retail environments, and healthcare spaces that people moved through during the day. Those spaces were organized around assumed occupants whose intersectional identities differed from those of the young sexual and gender minorities of color in this sample. Reaching that harm requires changing who institutional spaces are designed for and whose presence they assume, not only improving the neighborhoods surrounding them.

### 4.3 Implications for Intersectional Spatial Health Research

These findings carry specific implications for research and intervention oriented toward health equity for multiply marginalized populations. The three-mechanism architecture changes the questions that health equity intervention must ask. Current approaches typically ask whether material spatial conditions changed, whether a neighborhood gained a clinic, whether a workplace adopted a policy, or whether a park was built. SIHF requires a more precise question. Which mechanism was operating for which intersectional positions, and did the intervention address that mechanism for those specific intersectional formations? A greening initiative that improves material access to green space may still activate the Conditional mechanism for Black and Latina women if displacement follows. An affirming clinic that addresses cisgenderism may still activate the Conditional mechanism for queer people of color, producing harm along the racial axis that the nominally protective space leaves unaddressed. Non-binary people of color working under diversity policies designed around binary gender categories encounter the Positional mechanism in the same institutional space the policy was meant to address. Mechanism specification is the condition for reaching the source of harm rather than its most visible surface.

### 4.4 Limitations and Future Directions

The pilot sample of 19 GPS participants limits statistical power and generalizability, and adequately powered replication with samples large enough to include participants at contrasting intersectional positions navigating the same location types is the essential next step. The Positional mechanism’s core prediction, that the same space produces qualitatively different pathways for people at different intersectional positions, requires subgroup comparisons that pilot-scale designs can only approximate. The Layered mechanism had the lowest coded frequency in the pilot (5 of 91 records, or 5%). This reflects the pilot design rather than a null finding; the study was not powered to estimate mechanism-specific prevalence, and the compound measure’s consistent model superiority provides quantitative evidence consistent with Layered activation. The study was conducted in New York City, and how spatial intersectional mechanisms operate in smaller cities, suburban environments, and rural contexts remains an open question. A funded follow-on R01 study is designed to address these limitations directly, with a larger and more intersectionally diverse sample, revised sexuality structural exposure measures, and repeated monitoring across life circumstances. Returning to the account of Jasmine’s day, the central question addressed is not only who she is but whether the science designed to study her health was built to find the answer. Intersectionality had already named the interlocking systems targeting her. Spatial health research had already established that where she moved shaped her outcomes. What neither tradition had provided was an architecture for specifying exactly how those systems activate differently depending on which space she occupies, and what that does to her body in real time. That specification is what SIHF provides and what the methodology measures. The same space is not the same structural environment for everyone who enters it. The question now is whether health equity science is designed to act on that fact.

Jasmine opened this paper as a theoretical description. She closes it as an empirical one. The same space is not the same structural environment for everyone who enters it. SIHF and IGEMA provide the tools to show why, for whom, and through which mechanisms.

## 5. Conclusion

SIHF and IGEMA address a gap that neither intersectionality scholarship nor spatial health research could fill alone. The spatial architecture through which interlocking systems of oppression activate differently depending on who is moving through space. The three mechanisms specify that architecture. The GeoSense pilot demonstrates that it is empirically detectable. The field can now ask not only where health inequities cluster but also through which mechanism, for which identity formation, and at which specific places. Compound structural measurement outperforms single-axis alternatives. The positional mechanism accounts for most harm. Home proximity buffers acute stress for most participants while chronic structural disadvantage accumulates as baseline burden. These are not the same process, and they require different responses. Before SIHF and IGEMA, none of these distinctions was empirically accessible.

Interventions that address material spatial conditions without asking which mechanism is operating, and for which intersectional positions, will miss the source of harm for the communities most affected. Mechanism specification is not a refinement of existing approaches. It is the condition for designing research and intervention that actually reaches the structural architecture producing health inequity.

## Data Availability

All data produced in the present study are available upon reasonable request to the authors

