## Supplemental File for "Mapping Power Through Space: The Spatial Intersectionality Health Framework and Intersectional Geographically-Explicit Ecological Momentary Assessment"

**Table S2. Level 2 Multilevel Model Results: Full Output**

| **Predictor** | **b** | **SE** | **95% CI** | **p** | **d / LRT χ²** |
| --- | --- | --- | --- | --- | --- |
| **Panel A. Outcome: Mood (Mood_ND) — full analytic sample N = 32, 106 person-days** | | | | | |
| Within-person InDI (wp_indid) | -0.02 | 0.04 | [-0.10, 0.06] | .600 | d = -0.032 |
| Between-person InDI (bp_indid) | 0.01 | 0.08 | [-0.14, 0.16] | .879 | — |
| Marginal R² (discrimination-only) | — | — | — | — | R² = 0.033 |
| **Panel A continued. GPS subsample n = 15, 46 person-days — structural specifications** | | | | | |
| Racial + economic spec: LRT vs. Model A | — | — | — | .357 | χ²(4) = 4.4 |
| Pay gap spec: LRT vs. Model A | — | — | — | .112 | χ²(6) = 9.4 |
| ICE_gr spec: LRT vs. Model A | — | — | — | .007 | χ²(6) = 17.5 |
| wp_ice_gr | 26.75 | 7.21 | [7.54, 45.97] | .008 | — |
| bp_ice_gr | -27.62 | 8.68 | [-45.14, -10.09] | .003 | — |
| Marginal R² (ICE_gr model) | — | — | — | — | R² = 0.472 |
| **Panel B. Outcome: Perceived Stress (Stress_ND) — full analytic sample N = 32, 105 person-days** | | | | | |
| Within-person InDI (wp_indid) | 0.04 | 0.04 | [-0.04, 0.12] | .365 | — |
| Between-person InDI (bp_indid) | 0.13 | 0.05 | [0.04, 0.23] | .006 | — |
| Weekend (vs. weekday) | -0.27 | 0.13 | [-0.52, -0.02] | .037 | — |
| Marginal R² (discrimination-only) | — | — | — | — | R² = 0.032 |
| **Panel B continued. GPS subsample n = 15, 46 person-days — structural specifications** | | | | | |
| Racial + economic spec: LRT vs. Model A | — | — | — | .309 | χ²(4) = 4.8 |
| Pay gap spec: LRT vs. Model A | — | — | — | .426 | χ²(6) = 5.7 |
| ICE_gr spec: LRT vs. Model A | — | — | — | .004 | χ²(6) = 18.9 |
| wp_ice_gr | -33.33 | 9.86 | [-53.04, -13.63] | .002 | — |
| bp_ice_gr | 29.34 | 9.18 | [11.03, 47.65] | .003 | — |
| Marginal R² (ICE_gr model) | — | — | — | — | R² = 0.318 |
| **Panel C. Outcome: Sleep Disturbance (PROMIS) — full analytic sample N = 32, 134 person-days** | | | | | |
| Within-person InDI (wp_indid) | 1.34 | 0.46 | [0.43, 2.25] | .004 | d = 0.154 |
| Between-person InDI (bp_indid) | -0.08 | 0.72 | [-1.49, 1.33] | .912 | — |
| Marginal R² (discrimination-only) | — | — | — | — | R² = 0.021 |
| **Panel C continued. GPS subsample n = 12, 38 person-days — structural specifications** | | | | | |
| Racial + economic spec: LRT vs. Model A | — | — | — | .115 | χ²(4) = 7.4 |
| Pay gap spec: LRT vs. Model A | — | — | — | .287 | χ²(6) = 7.6 |
| ICE_gr spec: LRT vs. Model A | — | — | — | .094 | χ²(6) = 10.8 |
| **Panel D. Outcome: Physical Activity (METs) — full analytic sample N = 32, 132 person-days** | | | | | |
| Within-person InDI (wp_indid) | -0.01 | 0.01 | [-0.02, 0.00] | .087 | d = -0.105 |
| Between-person InDI (bp_indid) | 0.05 | 0.01 | [0.04, 0.07] | <.001 | — |
| Marginal R² (discrimination-only) | — | — | — | — | R² = 0.388 |

*Note. Models estimated using restricted maximum likelihood (REML) in R lme4/lmerTest. Full analytic sample models: three-level MLM (observations within days within persons), N = 32, n person-days varies by outcome. GPS subsample models: n varies by outcome (see panel headers). Predictor abbreviations: wp = within-person deviation; bp = between-person mean. b = unstandardized coefficient. SE = standard error. 95% CI = confidence interval. p values from Satterthwaite degrees of freedom. d = Cohen's d effect size. LRT = likelihood ratio test comparing Model B (+ structural exposure) vs. Model A (discrimination only). R² = marginal R² (Nakagawa & Schielzeth, 2013). InDI = Intersectional Denial and Discrimination Index (9-item nightly diary scale). ICE_gr = compound structural gendered racism index. — = not applicable or not estimated.*

*Physical activity (Panel D) within-person effect is from lagged model (wp_indid on Day D predicting METs on Day D+1; n = 24, 110 person-days). Between-person effect is from concurrent model.*

*Panel C GPS subsample sleep model did not reach significance threshold for LRT (p = .094); no individual structural coefficients reported for sleep ICE_gr model.*

**Table S4. Level 3 Coding Protocol and Intercoder Reliability**

| **Variable / Tier** | **N pairs** | **% Agreement (pre-cal)** | **κ (pre-cal)** | **% Agreement (post-cal)** | **κ (post-cal)** | **Notes** |
| --- | --- | --- | --- | --- | --- | --- |
| **space_type (OPPRESSION / RELIEF / COMPLEXITY / UNCLEAR)** | | | | | | |
| Tier 1 (location-specific) | 17 | 82% | 0.70 | 100% | 1.00 | Met pre-specified threshold (κ ≥ 0.70). Post-cal consensus on all 3 T1 disagreements. |
| Tier 2 (day-level narrative) | 48 | 79% | 0.68 | 100% | 1.00 | Marginally below threshold. 13 disagreements resolved in calibration; all OPPRESSION vs. COMPLEXITY boundary cases. |
| **spatial_mechanism (LAYERED / POSITIONAL / CONDITIONAL / unassigned)** | | | | | | |
| Tier 1 (location-specific) | 9 | 89% | — | — | — | Insufficient pairs for reliable kappa. 1 disagreement (ID 82, subway commute) resolved as Layered. |
| Tier 2 (day-level narrative) | 28 | 79% | 0.54 | 93% | 0.80 | Below threshold pre-cal. Disagreements at Layered-Conditional boundary. 2 records (IDs 65, 78) submitted for third-coder adjudication; resolved as Conditional. |

**Table S4b. Mechanism Count Summary (Primary Coder, n = 92 records, 18 participants)**

| **Mechanism** | **n (primary coder)** | **% of 92 records** | **Notes** |
| --- | --- | --- | --- |
| Positional | 66 | 72% | Dominant mechanism. Most harm occurred in institutional spaces regardless of structural disadvantage level of surrounding tract. |
| Conditional | 10 | 11% | Nominally affirming spaces with harm along unprotected identity dimension. |
| Layered | 5 | 5% | Compound multi-system activation at specific high-disadvantage locations. |
| Unassigned/UNCLEAR | 11 | 12% | Insufficient transcript content for mechanism assignment. |

*Note. Pre-cal = pre-calibration (independent first-pass coding). Post-cal = post-calibration (following structured calibration session). κ = Cohen’s kappa. Records coded UNCLEAR by either coder were excluded from kappa calculations per protocol. Calibration session: coders met to review all disagreements, discussed decision rules, and reached consensus on anchor cases. Third-coder adjudication: two mechanism records (IDs 65 and 78, Layered-Conditional boundary) reviewed by third coder; both resolved as Conditional. Tier 1 mechanism kappa not calculated due to insufficient pairs (n = 9). Primary coder = [removed for blind review]; secondary coder = [removed for blind review].*

**Supplement Note S1. IGEMA Level 3 Codebook: Classification Rules**

**Overview**

The IGEMA Level 3 Codebook (v7) governs assignment of qualitative codes to GPS-linked interview transcript excerpts. Each excerpt (Key Passage) is drawn from the map-guided qualitative interview conducted with participants after the monitoring period and linked to either a specific GPS dwell location (Tier 1) or a participant-day narrative (Tier 2). Coders assign two primary variables per record: space_type and spatial_mechanism. A third variable, intersection_active, identifies which intersecting systems of oppression were operating at the coded location.

The space_type categories adapt the Relief Maps framework (Rodo-de-Zarate, 2014). IGEMA uses the three categories as an intermediate analytic step within the SIF mechanism inference procedure, not as its endpoint. Full theoretical justification appears in the main text, Section 3.2.

**Two-Tier Coding Structure**

Tier 1 (T1; n = 22 records) comprises location-specific GPS dwell records where the transcript excerpt anchors to a named place. T1 records support place-level coding; both space_type and spatial_mechanism are assigned at the level of the individual GPS location.

Tier 2 (T2; n = 70 records) collapses shared_passage and general_day records into one row per participant-day. T2 records capture narrative spatial patterns across a day. Space_type and spatial_mechanism are assigned at the day-pattern level, not at a specific GPS location. The two-tier structure preserves analytical rigor by matching coding granularity to the evidentiary support available for each record type.

**Variable 1: space_type**

OPPRESSION. Assign when the Key Passage contains accounts of harm, surveillance, constraint, microaggression, or discrimination linked to the participant's intersectional positions in that space.

RELIEF. Assign when the Key Passage contains accounts of safety, belonging, reduced vigilance, or freedom from identity-based harm. A space can be stressful for reasons unrelated to structural power and still be coded RELIEF if identity-based harm is absent.

COMPLEXITY. Assign when OPPRESSION and RELIEF are simultaneously present and cannot be separated without losing analytical information. COMPLEXITY is not a residual code; it is correct when a space provides meaningful protection along one identity dimension while producing harm along another.

UNCLEAR. Assign when the Key Passage does not provide sufficient evidence for a substantive code. Blank cells are protocol violations; UNCLEAR indicates the coder evaluated the passage and determined evidence is insufficient.

**Variable 2: intersection_active**

intersection_active identifies which intersecting systems of oppression were operating at the coded location. SINGLE_AXIS codes are used when one primary system was activated. MULTI codes are used when two or more systems were simultaneously activated in an interlocked rather than additive way. intersection_active is not assigned when space_type is UNCLEAR.

**Variable 3: spatial_mechanism**

LAYERED. Multiple systems of oppression activate simultaneously at the location through the structural organization of the space itself. Convergence evidence: space_type is OPPRESSION or COMPLEXITY, and Level 1 structural exposure shows high simultaneous loading on racial and economic dimensions.

POSITIONAL. Harm follows the participant's intersectional position rather than the structural organization of the place. Divergence evidence: OPPRESSION or COMPLEXITY coded at a location where Level 1 structural data does not predict harm, or harm available across structurally opposite sites.

At Tier 2 (day-pattern level), Positional mechanism assignment is supported by convergent evidence of the same harm dynamic across structurally varied locations. When a participant describes encountering the same form of identity-based harm — being positioned as other relative to a space’s assumed occupant — across locations that differ in structural disadvantage profile, the cross-structural consistency confirms the Positional signature: harm travels with the person’s intersectional formation rather than concentrating where structural disadvantage is highest. A single location-level passage is not required; the day-pattern passage must capture a spatial logic in which the participant’s intersectional position, not the neighborhood’s structural composition, organizes the harm. This cross-structural pattern evidence is the primary inferential basis for Positional coding at T2, and it is often more legible at T2 than at T1 precisely because T2 passages capture the repeated pattern across multiple structurally varied locations rather than a single encounter.

CONDITIONAL. A nominally protective or affirming space that carries hidden costs at the participant's specific intersectional position. The harm operates through design assumptions about the expected occupant rather than aggregate structural disadvantage. COMPLEXITY is typically the correct space_type for Conditional sites.

**Triangulation Logic**

Level 3 codes are interpreted in relation to Level 1 structural exposure data and Level 2 multilevel model results. Convergence occurs when space_type and spatial_mechanism assignments are consistent with structural conditions at Level 1 and health effects at Level 2. Divergence, where assignments do not match Level 1 predictions, is theoretically informative rather than a coding error. Full triangulation procedures are described in the main text, Section 3.3, and illustrated in Section 3.3.4.

Full codebook text is available from the corresponding author on request.

**Supplement Note S2. GPS Data Processing Pipeline**

**Device and Export**

GPS data were collected via participants’ personal smartphones using Google Maps location history. Participants enabled location tracking throughout the monitoring period and shared their Google Maps Timeline data file with the study team at completion. Raw location data were processed to identify location visits (dwell events) using a dwell detection algorithm with a minimum dwell threshold of five minutes and a spatial radius of 50 meters.

**Place Name Assignment**

Raw GPS coordinates were matched to place names using two sources. Primary place name assignment used Google Maps Timeline data when available, which provides commercially enriched place labels for identified dwell locations. For records without Timeline matches, place names were assigned using OpenStreetMap (OSM) reverse geocoding via the Nominatim API with a maximum matching radius of 100 meters. OSM attributes (name, amenity type, shop type) were retained as structured fields. Records with no OSM match within 100 meters were assigned the resolved street address from Google Maps Geocoding API.

**Land Use Classification**

Each GPS visit record was classified using two sources of land use information. New York City PLUTO (Primary Land Use Tax Lot Output, v22.2) data were joined to GPS records by spatial intersection with tax lot polygons. PLUTO land use codes were used to assign residential, commercial, and mixed-use classifications at the parcel level. OSM amenity and shop tags provided place-type classifications for locations not covered by PLUTO (e.g., parks, transit stations, public spaces). A binary residential indicator was derived from PLUTO residential unit counts (is_residential_pluto = TRUE if UnitsRes > 0 and LandUse in residential categories).

**Dwell Time Calculation**

Dwell time at each location was calculated as the duration between the first and last GPS ping within the identified dwell cluster. Dwell events shorter than five minutes were excluded. Dwell events longer than eight hours were reviewed manually; extended dwell events at residential locations were retained as valid home visits. Extended dwell events at non-residential locations were flagged for review and retained if consistent with the participant's described schedule.

**Census Tract Assignment and Structural Indicator Linkage**

Each GPS visit record was assigned to a 2020 census tract using a point-in-polygon spatial join (R sf package, WGS84 coordinate reference system). Census tract GEOID was used to link American Community Survey (ACS) 5-year estimates (2017-2021) for all structural exposure indicators. Structural indicators were computed at the census tract level and merged to GPS records by GEOID. Person-level structural exposure values were computed as the time-weighted mean across all GPS visits for each participant on each monitored day, then averaged across all days to produce person-mean values.

**GPS Coverage and Exclusion Criteria**

Of 197 total GPS observation records, 169 were located within New York City borough boundaries (14 out-of-NYC records excluded from structural analyses). Records with no census tract match (n = 2) were excluded from Level 1 analyses. ICE_gr was available for 76 of 78 GPS-relevant census tracts (97.4% coverage); 2 tracts with missing ACS data were excluded from ICE_gr analyses only. Person-day structural exposure values were calculated from all available within-NYC records for each participant-day.

**Data Availability**

De-identified GPS coordinates and structural exposure values are available from the corresponding author upon reasonable request and subject to participant confidentiality protections. Participant home census tracts are not released to protect residential privacy.

**Supplement Note S3. Sexuality Structural Exposure: County-Level Limitation and Implications**

IGEMA's Level 1 architecture includes sexuality structural exposure as a third structural dimension alongside racial spatial polarization and economic deprivation. In this pilot, sexuality_struct_exp was operationalized using the proportion of same-sex households at the county level, derived from ACS 5-year estimates. This operationalization introduces a systematic limitation that affects the measure's utility for within-itinerary spatial analyses and warrants explicit documentation.

**Why County-Level Data Were Used**

Tract-level same-sex household counts from the ACS are unreliable in small-N samples, which characterize most New York City census tracts. The ACS uses a stratified sampling design that produces large margins of error for small-population subgroups at the tract level. In preliminary analyses, tract-level same-sex household proportions for the study area showed implausible variation and high rates of suppressed values, precluding their use as a spatially varying structural indicator. County-level aggregation (New York City borough level) was therefore used to produce a stable estimate.

**Consequences for the Pilot Analyses**

Because the sexuality_struct_exp variable is assigned at the county level, all participants residing and moving within the same borough receive the same value on every monitoring day. Within-person variation in sexuality_struct_exp across GPS visits is near-zero for participants who remained within a single borough, and between-person variation reflects only the borough of residence rather than variation in the structural sexual minority climate of specific visited tracts. Accordingly, sexuality_struct_exp contributes to between-person comparisons at Level 1 but cannot contribute to the within-person structural moderation analyses that are the primary focus of IGEMA's Level 2 layer. This measure is retained in descriptive reporting for completeness but is not included in the Level 2 structural specifications.

**Implications for Future Studies**

Adequately powered spatial analyses of sexuality structural exposure require tract-level indices that are not currently available from standard census products. Several alternatives have been proposed for future use. First, tract-level proportion of same-sex households using pooled multi-year ACS estimates with explicit uncertainty quantification. Second, community-based measures of sexual minority climate derived from participant-reported experiences of specific neighborhoods, following the spatial methods developed by Hatzenbuehler and colleagues (2014). Third, proprietary indices of anti-LGBTQ+ climate at the ZIP code or neighborhood level, where available. A funded follow-on R01 study is designed to incorporate multiple sexuality structural exposure indices at varying geographic resolutions to test the sensitivity of findings to operationalization choice. Until tract-level sexuality structural exposure can be reliably measured, IGEMA applications should treat sexuality structural exposure as a between-person contextual variable rather than a within-person spatially varying indicator.
